# Identification of Disease-Specific Turning Movement Hallmarks: A Systematic Review towards Establishment of Disease Screening Algorithm

**DOI:** 10.1101/2022.05.27.22275714

**Authors:** Ami Ogawa, Takanori Takeda, Kohei Yoshino, Hirotaka Iijima

## Abstract

**Background:** Patients with nervous system and musculoskeletal diseases display gait disturbance that is a leading cause of falls. Identification of disease-specific movement hallmarks is therefore an essential first step in preventing falls. Since turning, a common daily activity, is a unique movement that requires inter-limb spatial coordination, turning may be a suitable observational target for the identification of disease-specific movement disorder. However, to date, few comprehensive systematic review regarding disease-specific alterations in turning movement is available.

**Research question:** This systematic review with meta-analysis summarized the level of knowledge regarding movement disorders during turning in patients with nervous system and musculoskeletal diseases.

**Methods:** A systematic review was conducted of papers throughout 2021 in accordance with PRISMA guideline. Including criteria were (1) were published in a peer-reviewed journal, (2) were written in English, (3) included adult patients who were diagnosed with musculoskeletal or nervous system diseases, (4) had a control group of age-matched healthy adults, and (5) outcomes included turning parameters.

**Results:** Meta-analysis revealed a significantly larger step number, longer turn duration, and shorter step length in patients with Parkinson’s disease (PD) than in controls during the 180° turn, suggesting that these biomechanical alterations may be, at least in part, movement disorders associated with PD. Notably, this review identified methodological heterogeneity for turning movement assessments, which limited the identification of disease-specific movement disorders.

**Significance:** This work serves as a call to action for the establishment of a standard assessment protocol towards the identification of disease-specific turning movement disorders and effective disease screening.

## Introduction

Diseases with gait disturbances, such as nervous system and musculoskeletal diseases, present a high risk of falling in patients, because of limitation of joint movement or lack of balance ability. For instance, in patients with Parkinson’s disease (PD), a symptom called freezing of gait (FOG), which causes the sudden loss of the ability to start or continue walking[1], is known as one of the causes of falls[2]. Patients with stroke also have a high risk of falls because of a decline in physical abilities[3]. Preventing falls is required due to the high mortality[4]. Detecting disease-specific early hallmarks is important in delaying disease progression and preventing falls; thus, enabling the appropriate intervention for each disease in the early stages.

Identification of movement disorder during daily activities is a potential staple for the disease early detection. Among daily activities, turning accounts for 8–50% of steps taken within a day[5] and is more physically challenging than level walking that requires inter-limb spatial coordination[6, 7]. Since studies has successfully characterized turning movement disorder in several neurological and musculoskeletal diseases including PD[8, 9] and stroke[10, 11], turning may be a suitable observational target for the disease screening. Identifying disease-specific movement disorder during turning is particularly important given that falls during turning is 7.9 times more likely to cause a hip fracture than those from straight walking in the elderly people[12].

Systematic review with meta-analysis of the literature allows for comparison of inconclusive results across different studies and identification of turning movement hallmarks associated with disease. Although, previous systematic review characterized movement disorder associated with PD[13], no systematic review summarized turning characteristics across a wide range of diseases that is an essential next step towards the identification disease-specific movement disorder and subsequent disease screening. With this in mind, this systematic review with meta-analysis aimed to summarize level of knowledge regarding the turning movement characteristics in patients with nervous system and musculoskeletal diseases with the goal of identification of disease-specific turning movement disorder. Through this review, we identified methodological heterogeneity for turning movement assessment, which builds a framework for future biomechanical studies.

## Methods

This study was conducted according to the Preferred Reporting Items for Systematic reviews and Meta-Analyses (PRISMA) statement[14], PRISMA protocols (PRISMA-P)[15], Meta-analysis of Observational Studies in Epidemiology (MOOSE) checklist[16], and Cochrane Handbook for Systematic Reviews of Interventions[17].

### Literature search and study selection

PubMed, Physiotherapy Evidence Database, Cumulative Index to Nursing and Allied Health Literature, and Cochrane Central Register of Controlled Trials were searched. Included studies met the following criteria: (1) were published in a peer-reviewed journal, (2) were written in English, (3) included adult patients who were diagnosed with musculoskeletal or nervous system diseases, (4) had a control group of age-matched healthy adults, and (5) outcomes included turning parameters. We defined the turn movements as a change of direction to a specific angle before or after a straight walk; thus, a standing turn (turning on the spot) was also included. Diagnostic criteria for musculoskeletal or nervous system diseases were defined by each study’s inclusion criteria. Since turning parameter changes during aging, we included only studies with an age-matched control group. No restrictions on study dates, follow-up duration, disease severity, or time since disease onset were used. Electronic searches used combined key terms of PubMed, including “Musculoskeletal Diseases,” “Nervous System Diseases,” “Turning,” “Turn,” “Biomechanical Phenomena,” “Kinetics,” and “Accelerometry” using Medical Subject Headings terms.

In the first review, a single reviewer (HI) conducted an electronic database search throughout June 2019. Two independent reviewers (TT and KY) assessed eligibility in a blinded manner in accordance with the Cochran Handbook recommendations[17]. The second review was conducted by two independent reviewers (AO and TT) throughout April 2021 to ensure that further articles were also assessed for inclusion prior to publication. After duplicates were removed, the two reviewers screened titles and abstracts yielded by the database search. Full manuscripts of the articles that met the eligibility criteria were then obtained and reviewed. During these processes, the reviewers prepared and used simple, pre-designed Google spreadsheets to assess eligibility by extracting study features. Disagreements between the two reviewers were discussed until consensus was achieved.

### Outcome measure and data extraction

The primary outcomes in this review were (1) spatiotemporal parameter (i.e., turn velocity, number of steps), (2) kinematics (i.e., joint angle, foot clearance), (3) strategy parameters during a turn. The secondary outcomes were (4) electromyography (EMG) and (5) kinetics parameters during a turn.

Turning involves first recognizing the movement direction and then reorienting each segment and moving toward the new direction by stepping. The variations of movement are made when the new direction is recognized and how each segment is moved, i.e., turning strategy, because turning is accomplished over several steps. Thereby, turning strategy is accounted as one of the categories of outcomes in addition to the traditional outcome categories such as spatiotemporal, kinetics, kinematics, and EMG parameters. The word “strategy” is often used without any definition in previous studies. Thus, we originally defined the category of strategy in this review that includes following three subcategories: (1) how to step toward the new direction, i.e., turn type subcategory (few-step [spin turn and step turn] or multi-step turns), (2) spatial subcategory such as the trajectory of the center of mass (COM) or distance of COM respect to center of turn, and (3) segmental subcategory such as the delay time of reorientation of each segment that shows the linkage of each segment (head, trunk, and pelvis). Regarding turn types, few-step turns including spin and step turns were identified in which the number of steps from the start to the end of the turn is less than three, and those in which the number of steps exceeded four were distinguished as multi-step turn. The spin turn was defined as a strategy in which the foot ipsilateral to the turning direction is the axle foot and the opposite foot crosses the axle foot, while step turn was defined as a strategy when the foot contralateral to the turning direction is the axle foot and throw the ipsilateral side to the turning direction without crossing[18]. The overview of outcome categorization is shown in **Figure 1**.

**Figure 1.**
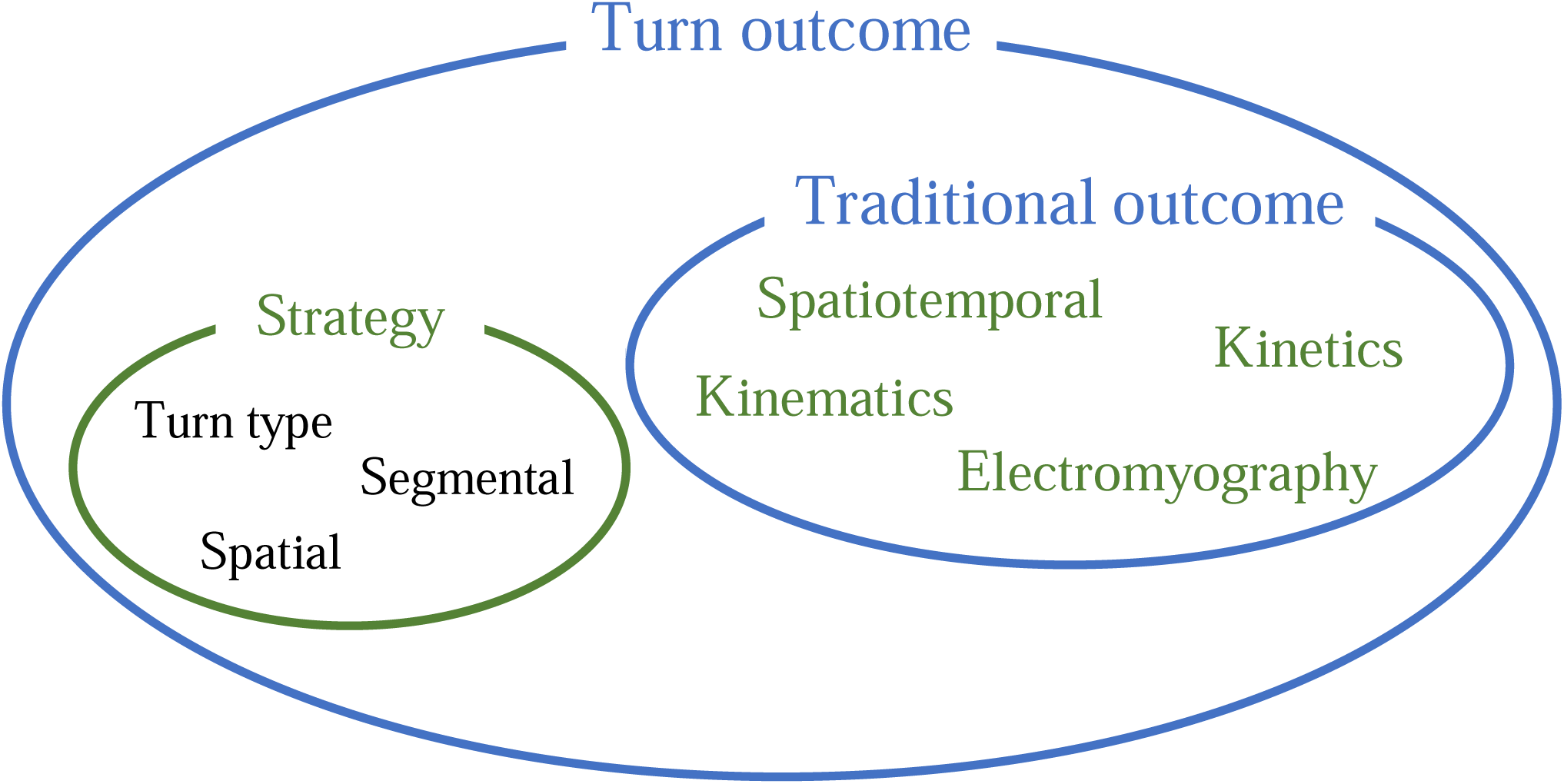
Structure of outcome categorization.

The outcome measures were categorized according to turning angle (90°, 120°, 180°, et al.) and whether it was a walking turn or standing turn. Turning angle is one of the factors that determine the turn strategies[8] and it makes the movements change; thus, data should be separately analyzed. Similarly, walking turn and standing turn require different movement mechanisms, and normally they were dealt with separately[19, 20].

Three reviewers (TT, KY, and AO) collected data regarding authors, publication years, subject population, subject disease, outcome measures, and turning tasks using standardized data forms. When mean and standard deviation (SD) values were not directly reported, they were calculated from other available data whenever possible, such as interquartile range[21] or we directly contacted the authors. When data were only provided in figures, the graphical data were converted to numerical data using a digital ruler software (WebPlotDigitizer 4.4)[22].

### Data Analysis

Meta-analyses were performed using Review Manager Version 5.4 (Nordic Cochrane Centre, Cochrane Collaboration, Copenhagen, Denmark). Pooled estimates and 95% confidence intervals for standardized mean differences (SMD) of outcome measures were calculated using a random effect model[23]. Study heterogeneity, the inter-trial variation in study outcomes, was assessed using *I*^*2*^, which is the proportion of total variance explained by inter-trial heterogeneity[24]. Meta-analyses were done for more than two studies that were gathered in each categorization on turning angle and whether it was a walking turn or standing turn. Data that were not considered appropriate for meta-analysis were also summarized in the table to conduct narrative synthesis.

## Results

The database search yielded a total of 735 studies; 53[8-11, 19, 20, 25-71] ultimately met the eligibility criteria and were included in the systematic review (**Figure 2**). The rationales for the exclusion of studies during the full-text screening process were incorrect outcome (47%), without the control group (21%), non-peer review journal (16%), and non-musculoskeletal or nervous system diseases (16%).

**Figure 2.**
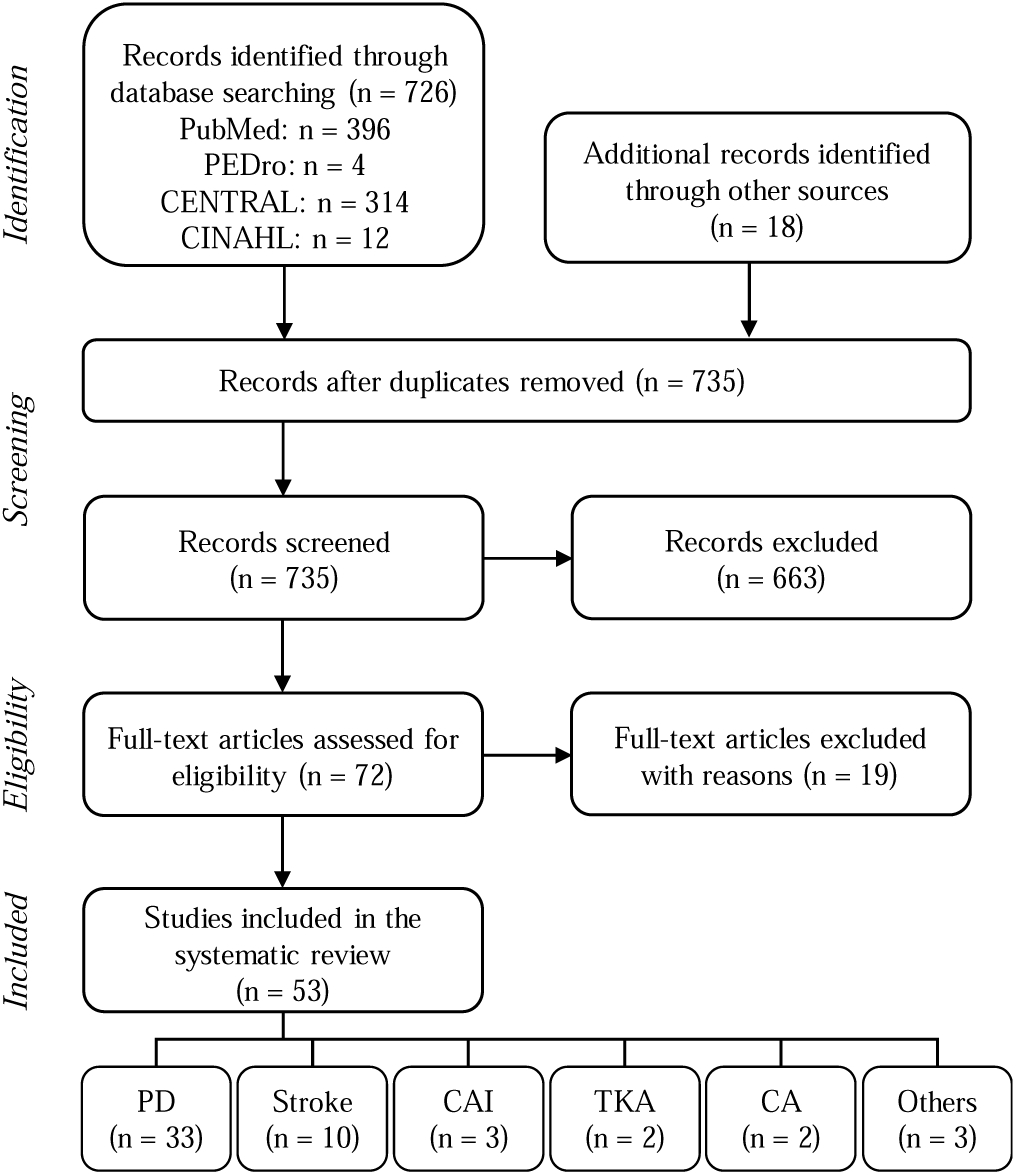
Flowchart of study selection according to Preferred Reporting Items for Systematic Reviews and Meta-analyses (PRISMA).

**Table 1** summarizes the characteristics of included studies. Most of the studies were on PD (33 studies) and stroke (10 studies) among nervous system diseases. The other studies included were on cerebellar ataxia (CA; two studies), Fragile X-associated tremor/ataxia syndrome (FXTAS; one study), and Huntington’s disease (HD; one study) among those categorized as nervous system diseases. Chronic ankle instability (CAI; three studies), total knee arthroplasty (TKA; two studies), and transmetatarsal amputation (TMA; one study) were categorized in musculoskeletal diseases. In the PD study, a total of 753 patients (mean age: 66.4 years) and 462 age-matched healthy adults (mean age: 67.3 years) were included from 33 studies. Regarding the stroke study, a total of 166 patients (mean age: 59.9 years) and 148 age-matched healthy adults (mean age: 52.4 years) were obtained from 10 studies.

**Table 1.**
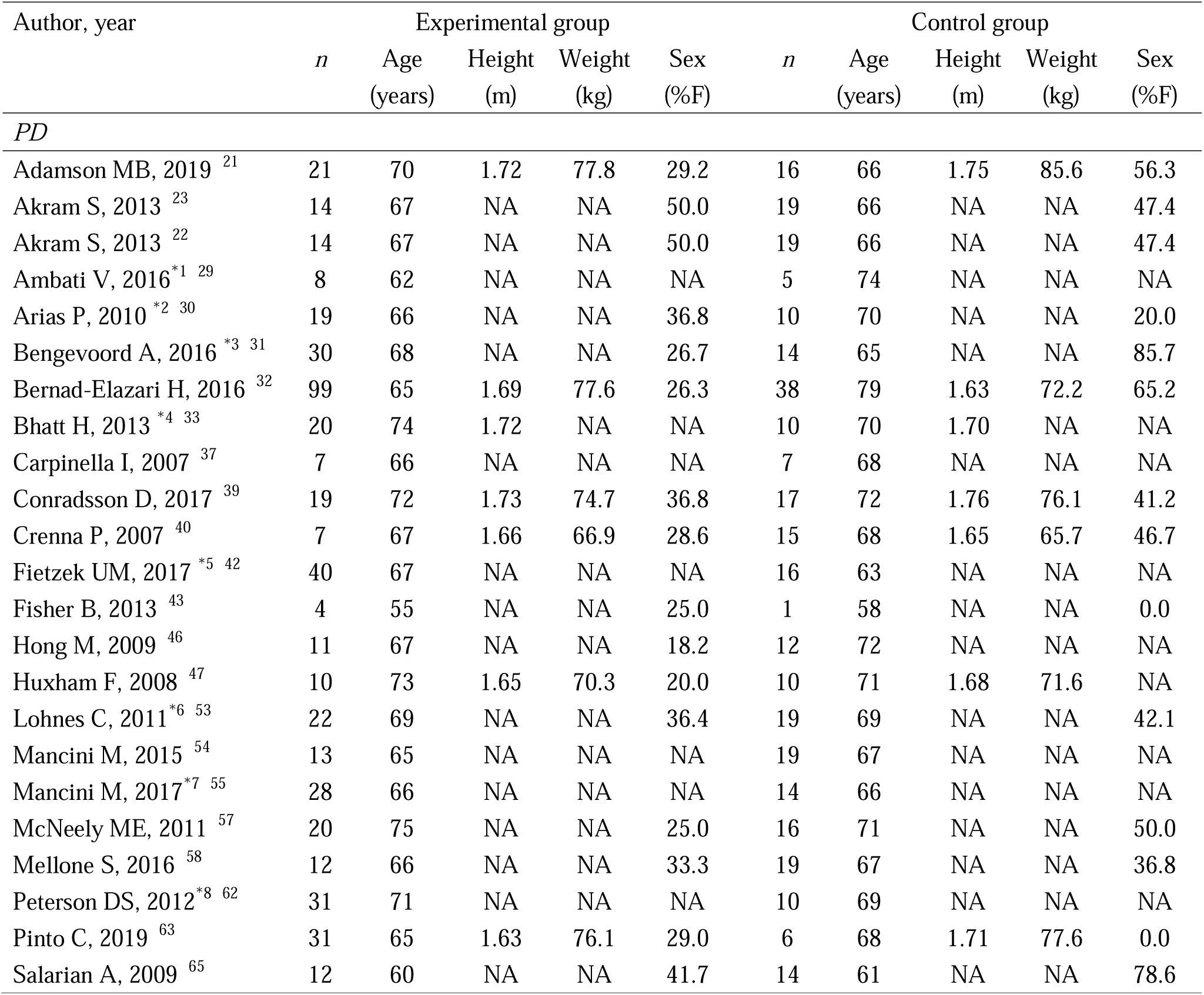
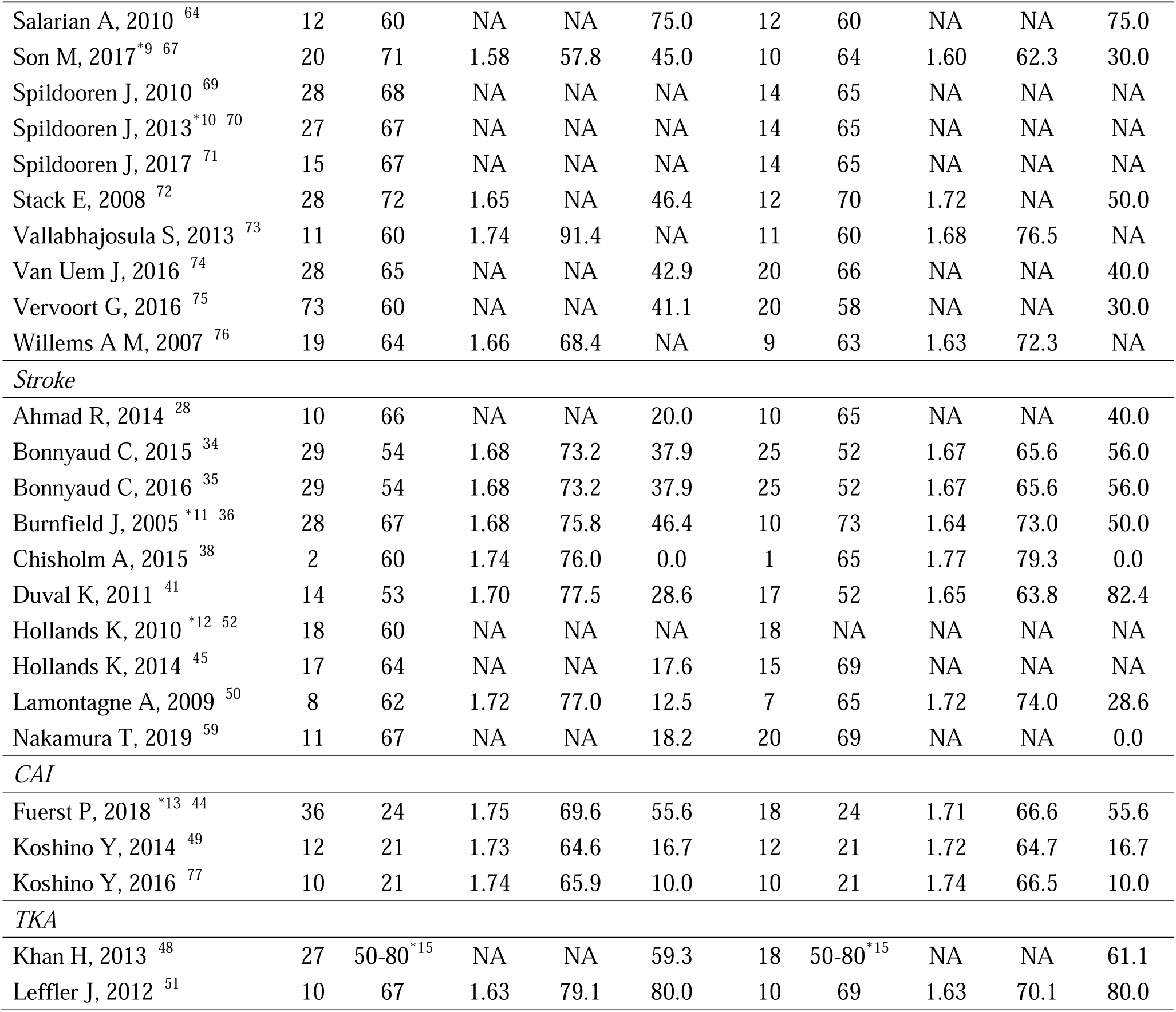
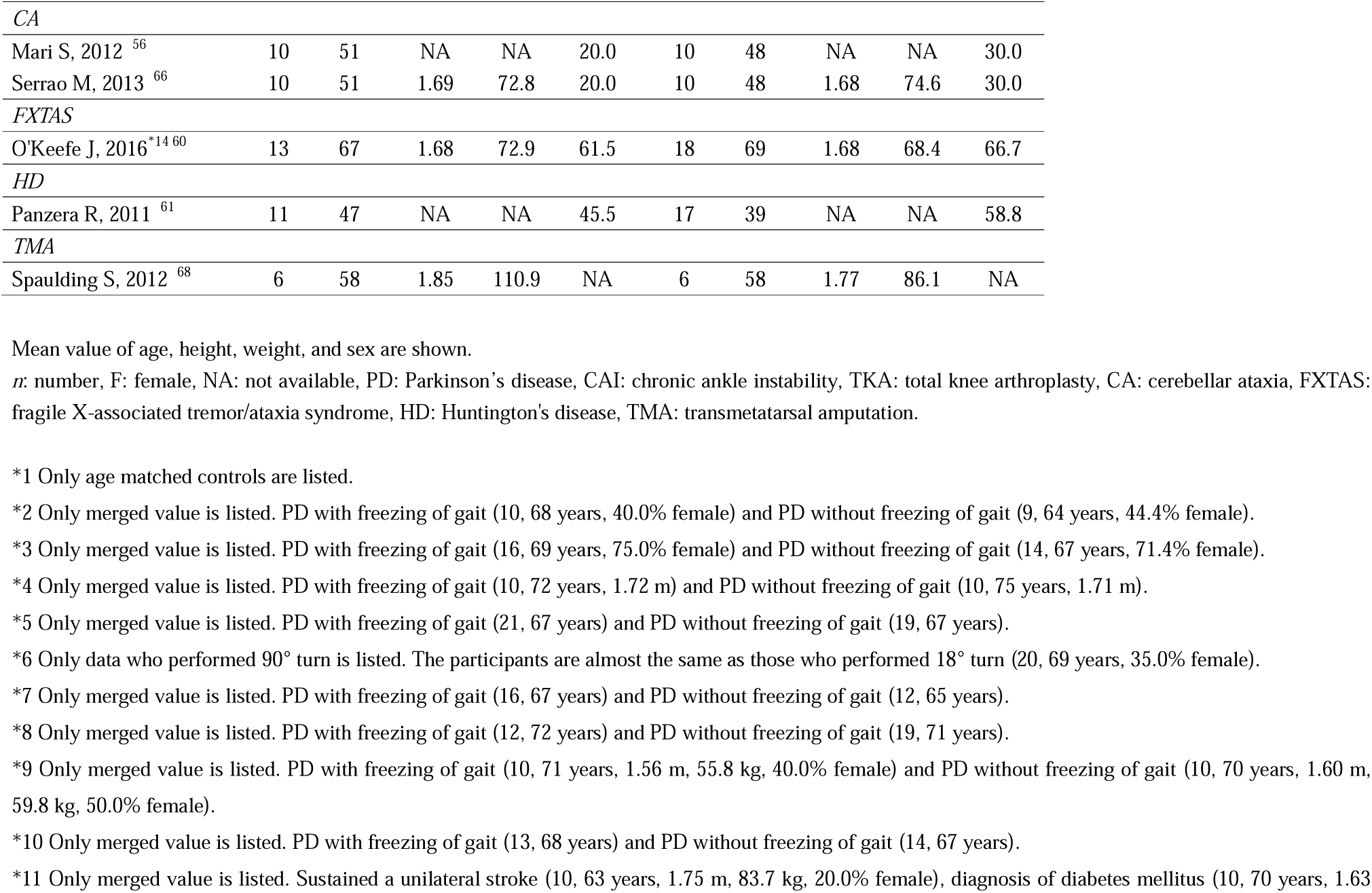
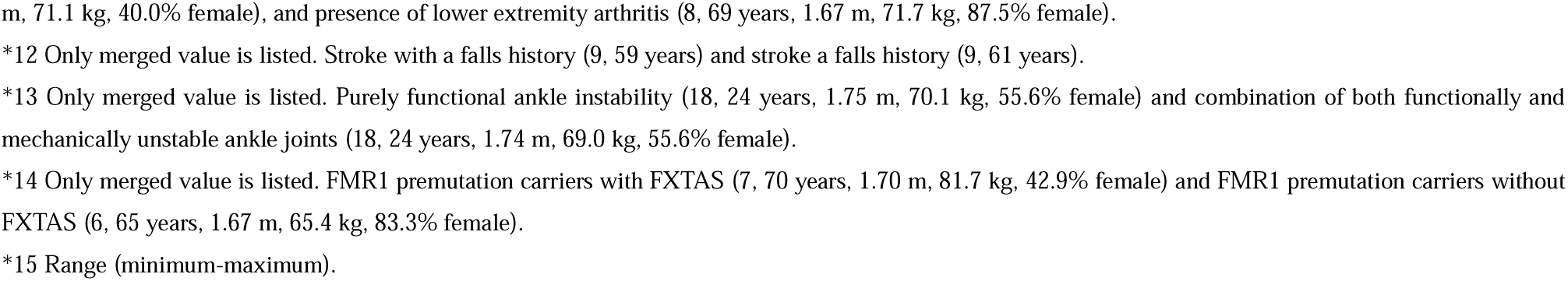
Characteristics of the included studies.

The outcomes and apparatus are shown in **Table 2**. The majority of apparatus used for turning analysis were: 1) three-dimensional motion capture system, 2) force plate, and 3) inertial measurement units. Surface EMG, eye tracker, footswitch, and video cameras were also used.

**Table 2.**
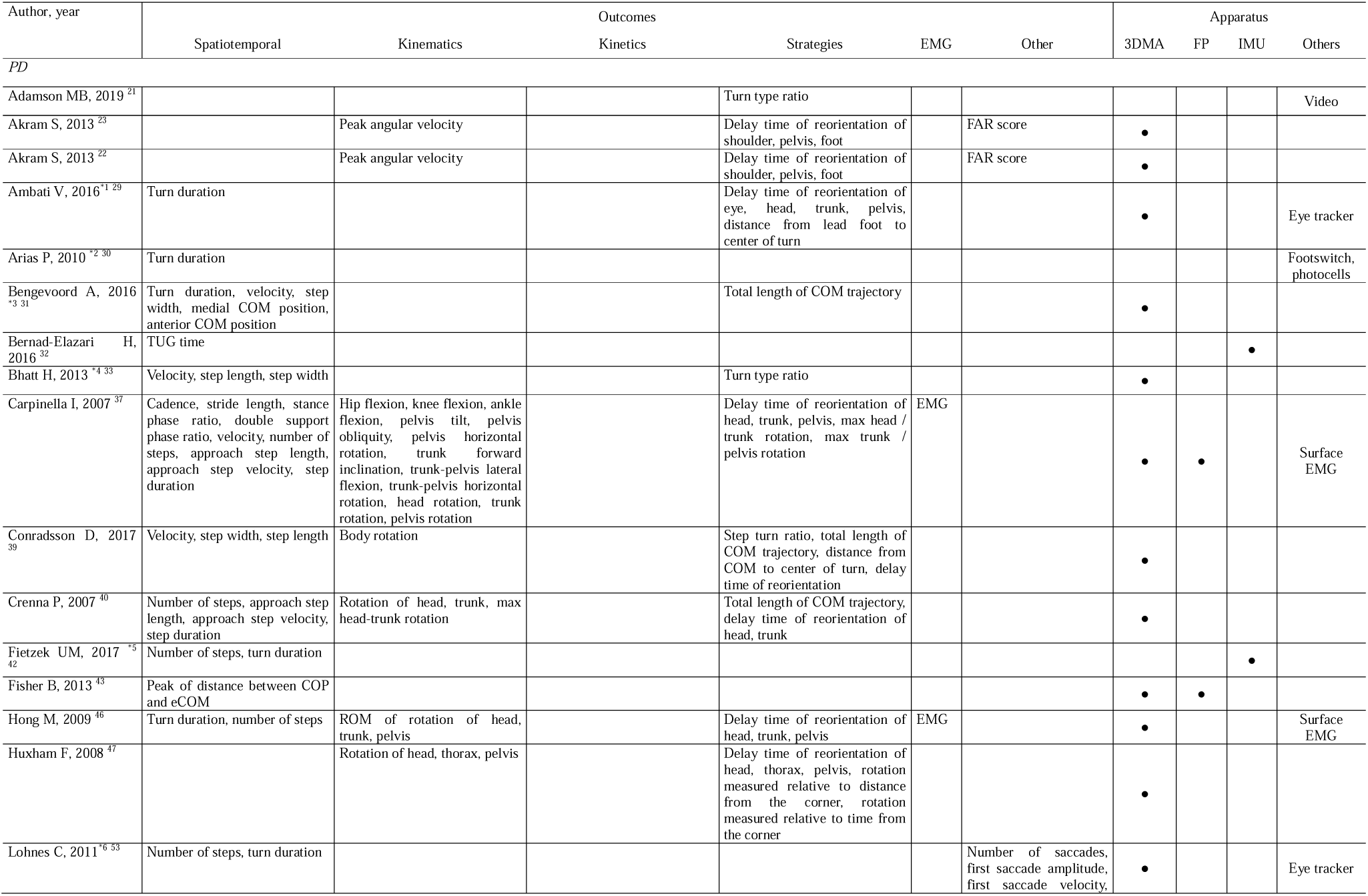
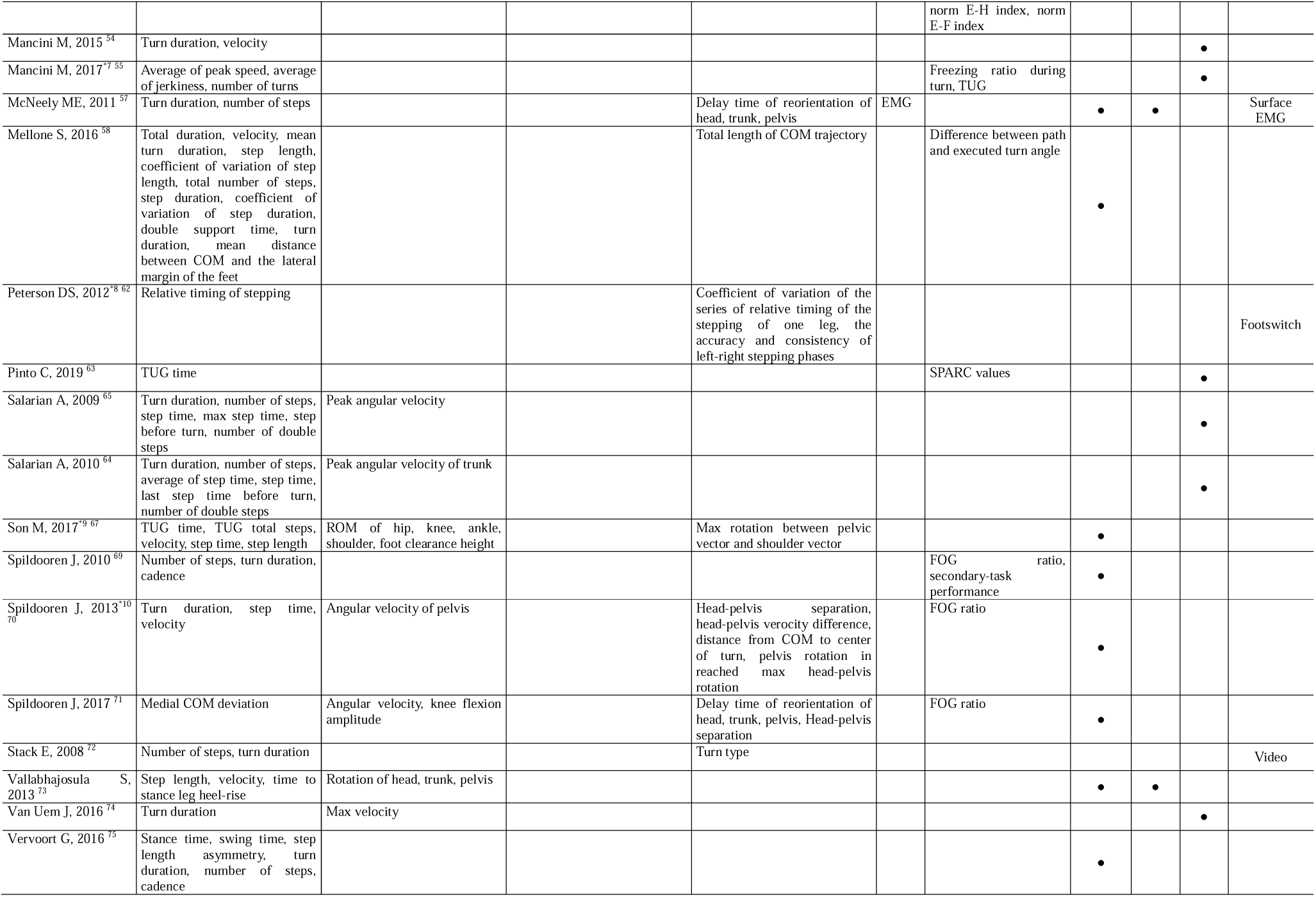
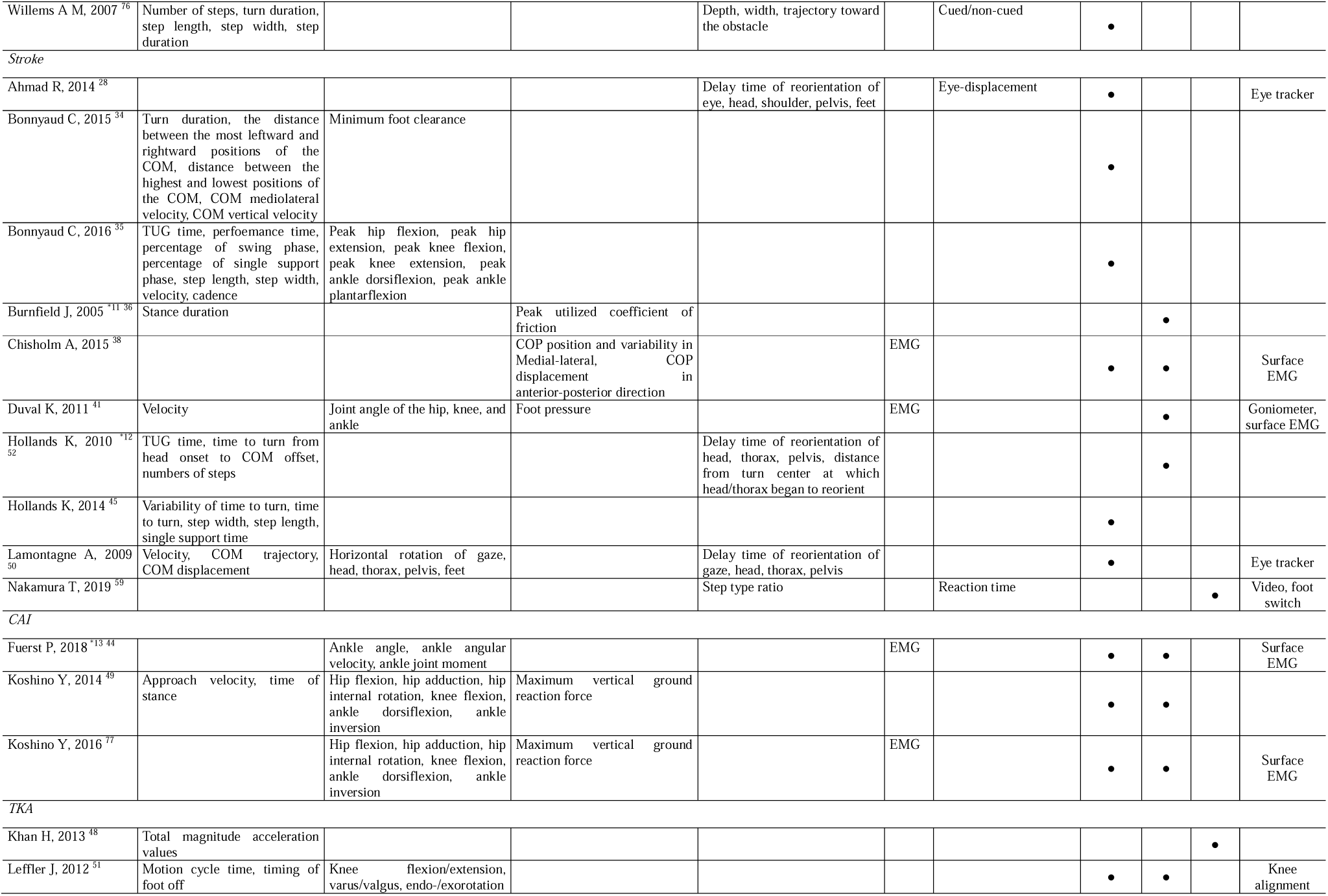
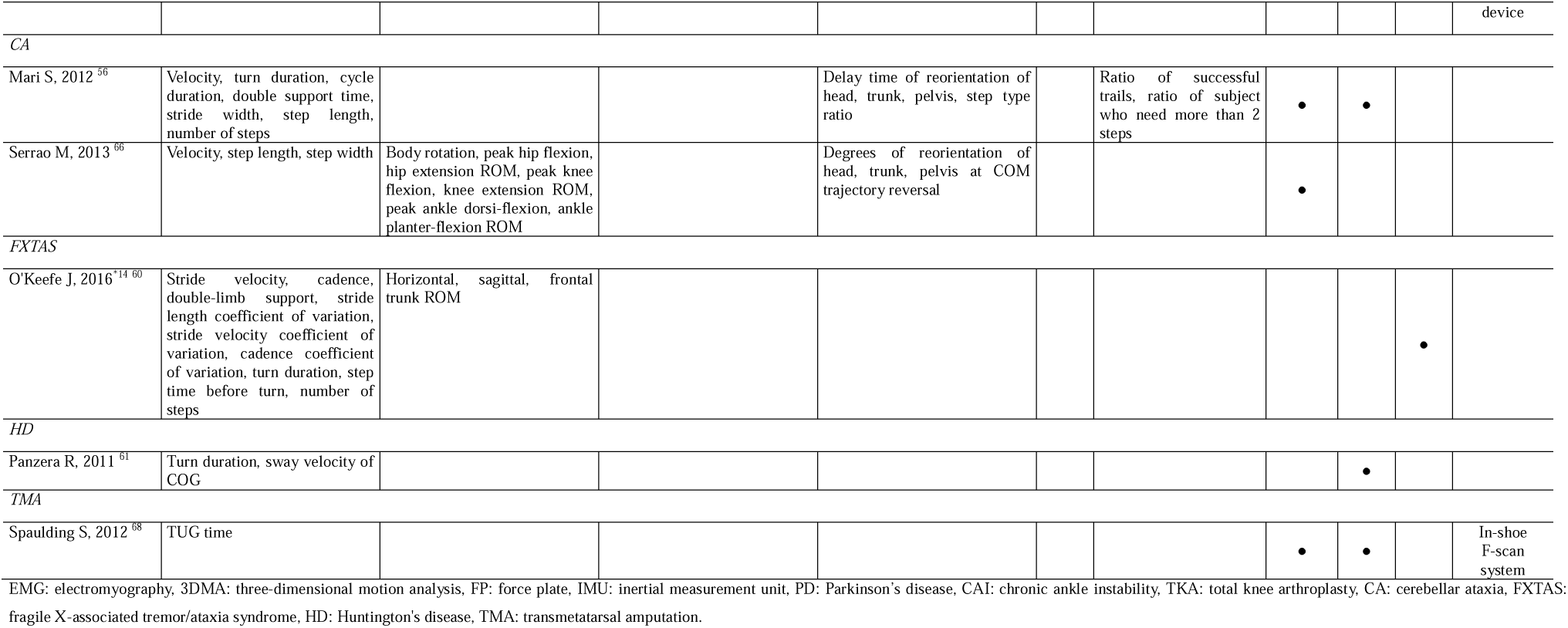
Outcomes and apparatus of included studies.

### Systematic review identified methodological heterogeneity for turning movements assessment

The conditions of turning tasks are shown in **Table 3**. An inconsistent method of turning task was revealed. The most common angle of turning was 180° in 31 cases, followed by 90° in 20 cases, 45° in 6 cases, 360° in 4 cases, 120° and 135° in 2 cases each, and 30° and 60° in 1 case each.

**Table 3.**
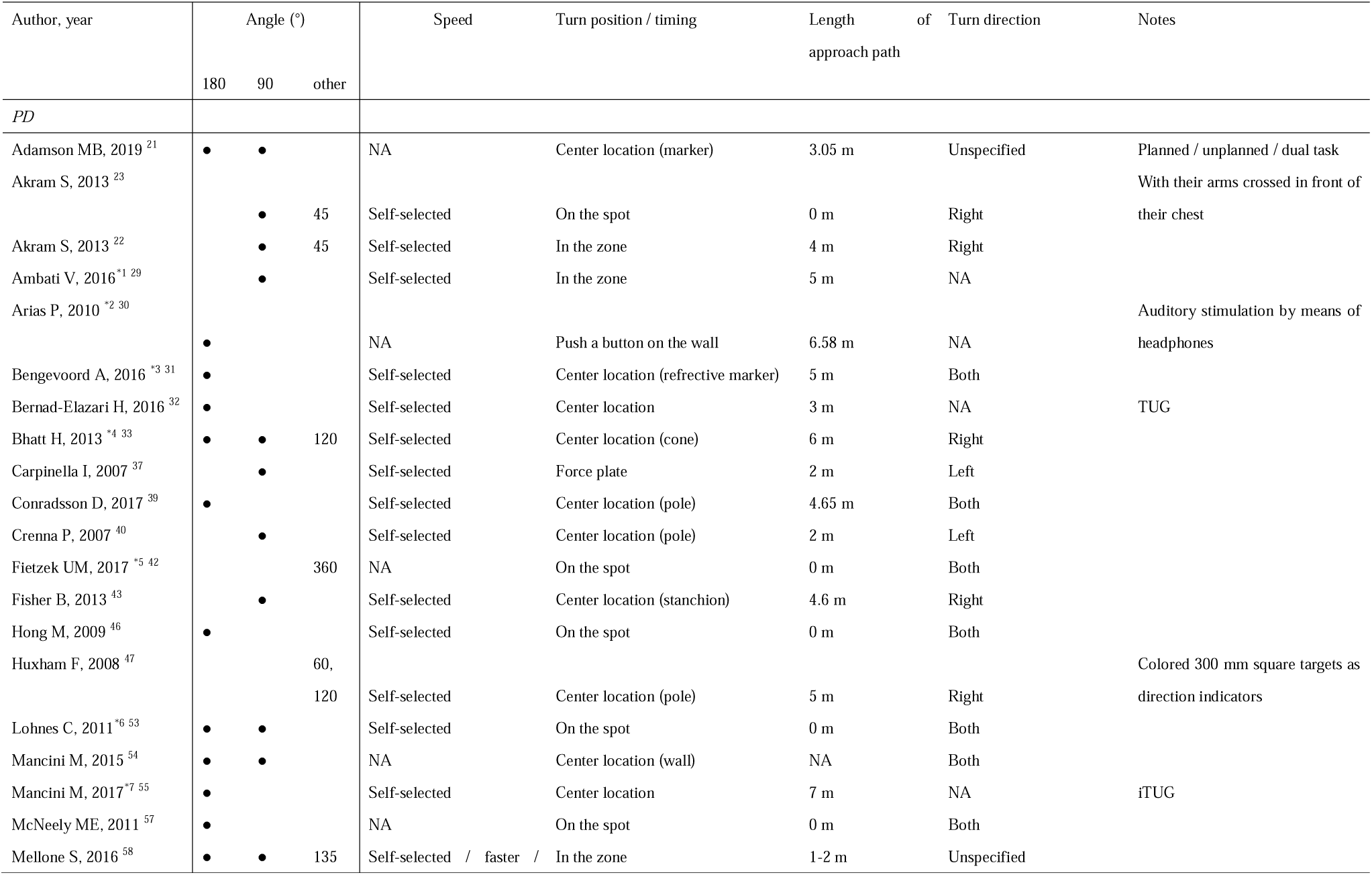
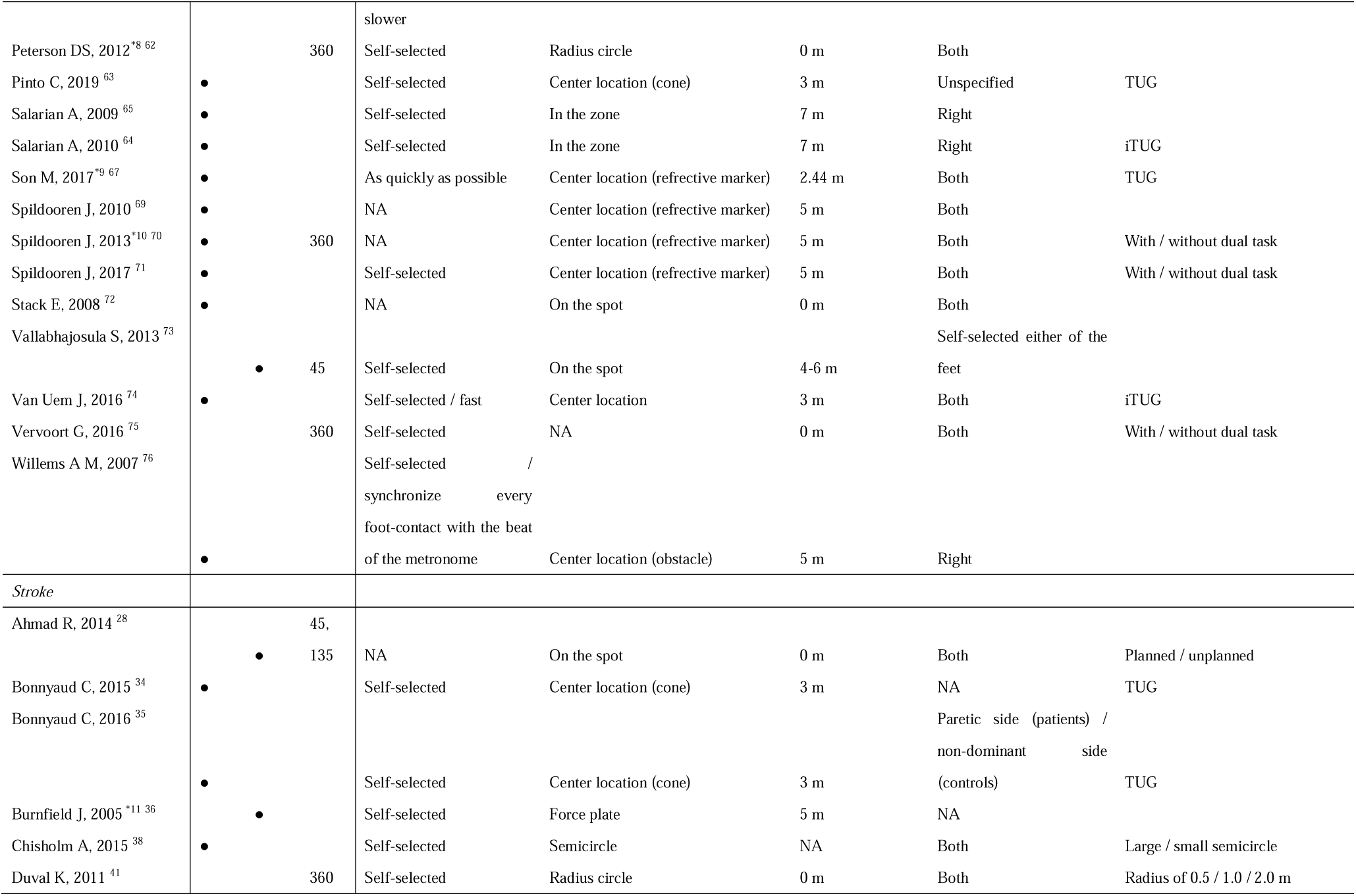
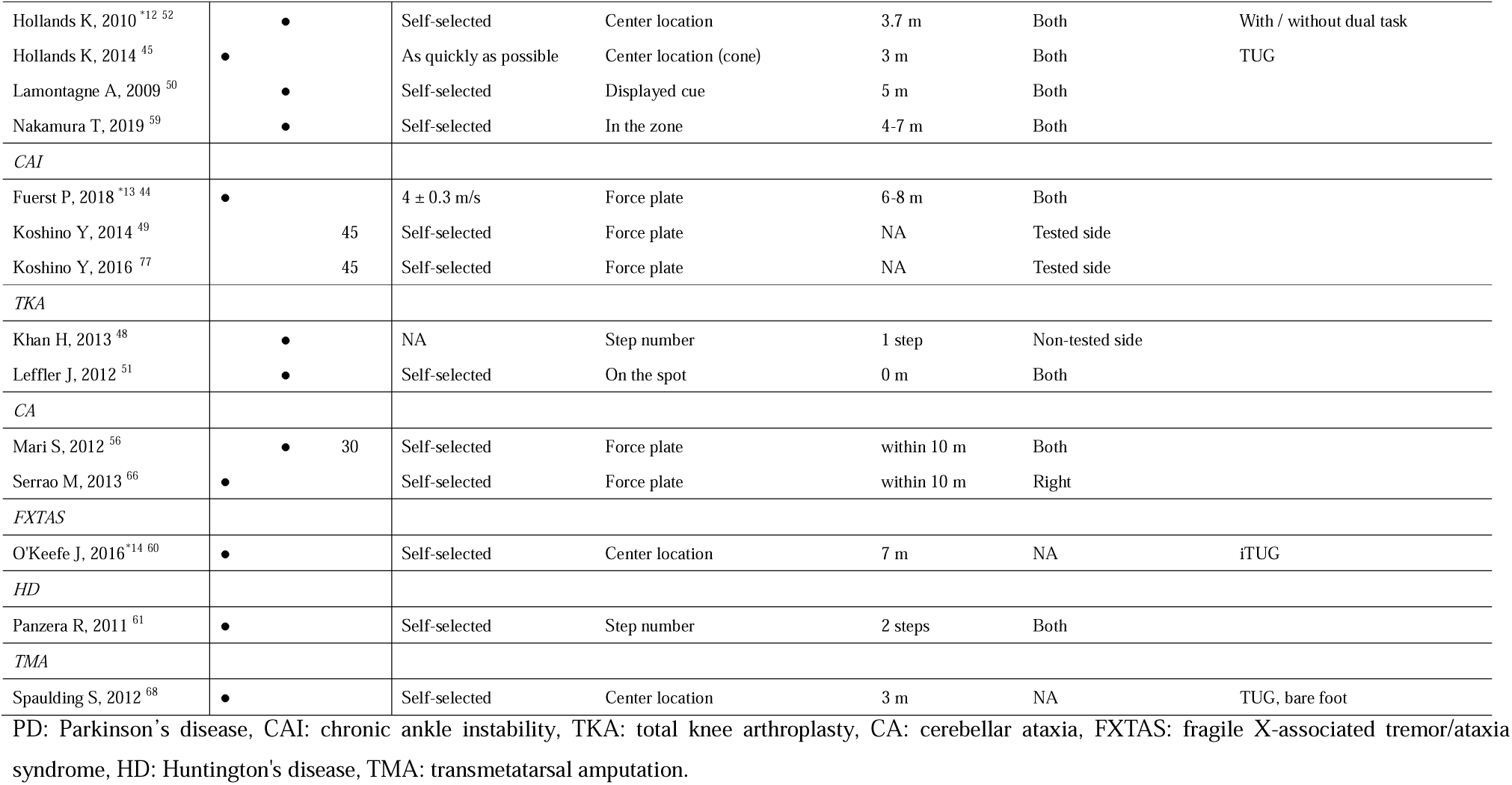
Turning conditions.

In most studies, participants were instructed to turn at a self-selected speed. In some cases, controls were instructed to walk at a slower speed to suit patients, and in other cases, several levels of speed were set (fast, comfortable, slow) or speed was specified.

In 24 studies[8, 9, 11, 25, 27-30, 34, 35, 38, 40, 42, 47, 49, 50, 55, 58, 62-65, 68, 70], the tasks were set the turning position with the center locations marked by cones, poles, stanchions, reflective markers, or tapes. Seven studies[31, 32, 39, 44, 51, 61, 71] defined the foot position of turning with a force plate. Five studies[19, 53, 54, 59, 60] defined a certain zone for a turn, and two studies[43, 56] instructed several steps before turning.

Nine studies measured turning on the spot[10, 20, 37, 41, 46, 48, 52, 66, 67], three studies were radius circle or semicircle[33, 36, 57]. The maximum and average length of approach path was 7 m and 3.33 m, respectively. Turn direction was set for both sides in 26 studies[9, 10, 27, 33, 34, 36, 37, 39-41, 45-49, 51, 52, 54, 56, 57, 62, 64-66, 68, 69], and 16 studies limited either side[11, 19, 20, 29, 32, 35, 38, 42-44, 59-61, 67, 70, 71]. The formalized task was only TUG with 7 studies[11, 28, 30, 40, 58, 62, 63], 11 studies including instrumented TUG[50, 55, 59, 68].

### Synthesis of Results

The summary of the meta-analysis is shown in **Table 4**. Only PD data was applied to meta-analysis, because of no outcomes commonly reported in more than one study or discrepancy of conditions for other diseases.

**Table 4.** Summary of meta-analysis. Dark blue cells: parameters that meta-analysis was performed. Alphabets are corresponding to Figure 3. -: data available but not enough study for meta-analysis because of discrepancy of conditions. PD: Parkinson’s disease, Walking: walking turn, Spot: walking on the spot.

|  |  | PD |  |  |  | Stroke |  |  |  |
| --- | --- | --- | --- | --- | --- | --- | --- | --- | --- |
|  |  | 90° |  | 180° |  | 90° |  | 180° |  |
|  |  | Walking | Spot | Walking | Spot | Walking | Spot | Walking | Spot |
| Spatiotemporal | Number of steps | - | - | A | A |  |  |  |  |
|  | Turn duration [s] |  | - | B | B | - |  | - |  |
|  | Average of step duration [s] | - |  | C |  |  |  |  |  |
|  | Turn velocity [m/s] |  |  | D |  |  |  |  |  |
|  | Step length [m] |  |  | E |  | - |  | - |  |
|  | Step width [m] |  |  | F |  | - |  | - |  |
| Kinematics | Angular velocity [°/s] |  |  | G |  |  |  |  |  |
|  | Head rotation [°] | - |  |  |  |  |  |  |  |
| Strategy | Step turn ratio [%] |  |  | - |  |  |  |  |  |
|  | Total length of COM trajectory [m] |  |  | - |  |  |  |  |  |
|  | Distance from COM to center of turn [m] |  |  | H |  |  |  |  |  |
|  | Delay time of pelvis reorientation [s] | I |  |  |  |  |  |  |  |
|  | Head-pelvis rotation [°] |  |  | J |  |  |  |  |  |

#### Patients with PD performed slow multi-step and “en bloc” turns

**Figure 3*A*** shows that the number of steps in PD significantly increased compared to the control group during the 180° turning[60, 64, 70] and 180° turning on the spot[41, 48, 52, 66]. A similar tendency was shown during 90° turning[32, 35] and 90° turning on the spot[48]. Turn duration in patients with PD was significantly longer than controls during 180° turning[9, 60, 64, 68, 70] and 180° turning on the spot[41, 48, 52, 66] (**Figure 3*B***). The duration during the 90° turning on the spot[48] showed a similar trend. There was a small differences in step duration between PD and control groups during 180° turning[9, 60, 62, 64, 70] (**Figure 3*C***), slightly longer in patients with PD in 90° turning[32, 35]. In addition, maximum step duration in the PD group during 180° turning was barely longer than controls[59, 60]. **Figure 3*D*** shows that turn velocity in patients with PD was slower than controls during 180° turning[29, 34, 62]. Similarly, the peak angular velocity of the trunk in the horizontal plane in patients with PD was smaller than controls during 180° turning in TUG[60, 68] (**Figure 3*E***). According to **Figures 3*F* and 3*G***, the step length[29, 62, 70] and step width[29, 70] are shorter in patients with PD during 180° turning.

**Figure 3.**
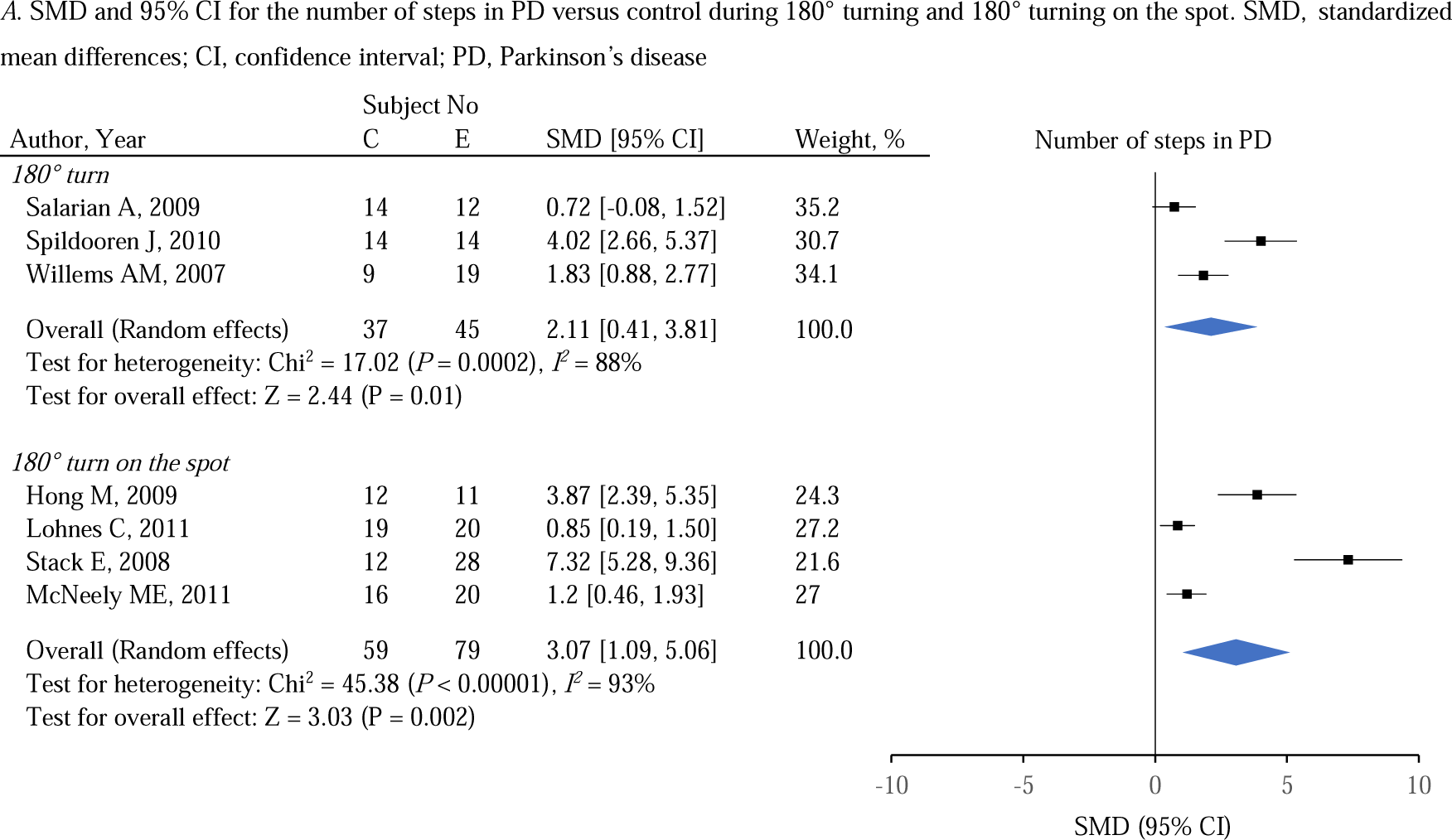
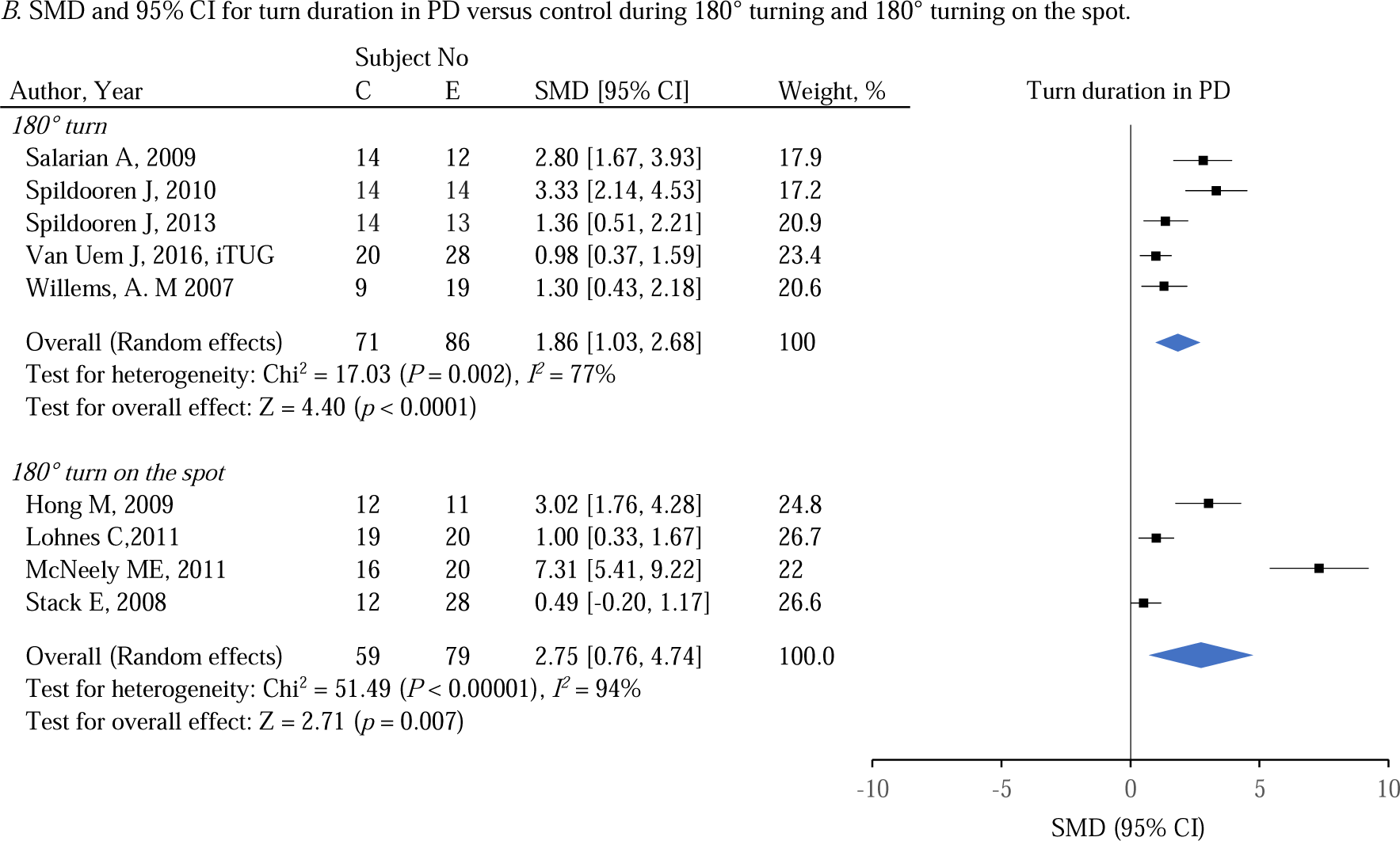
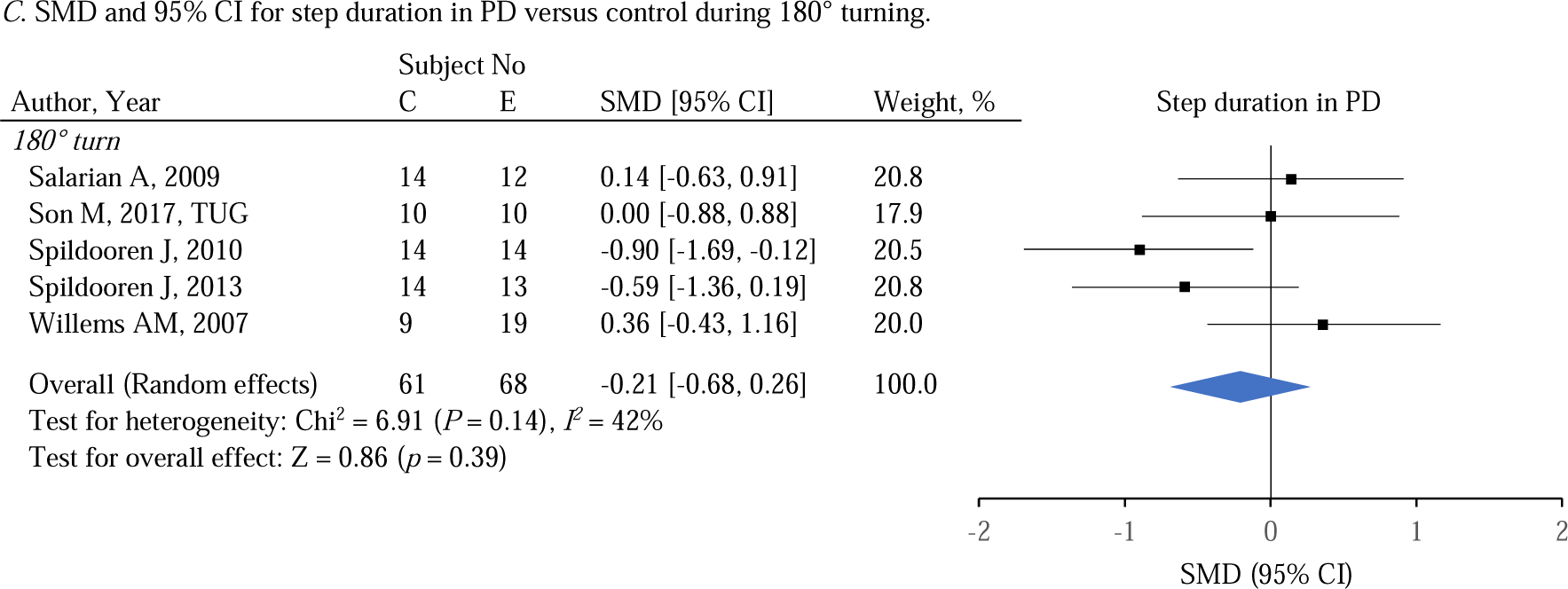
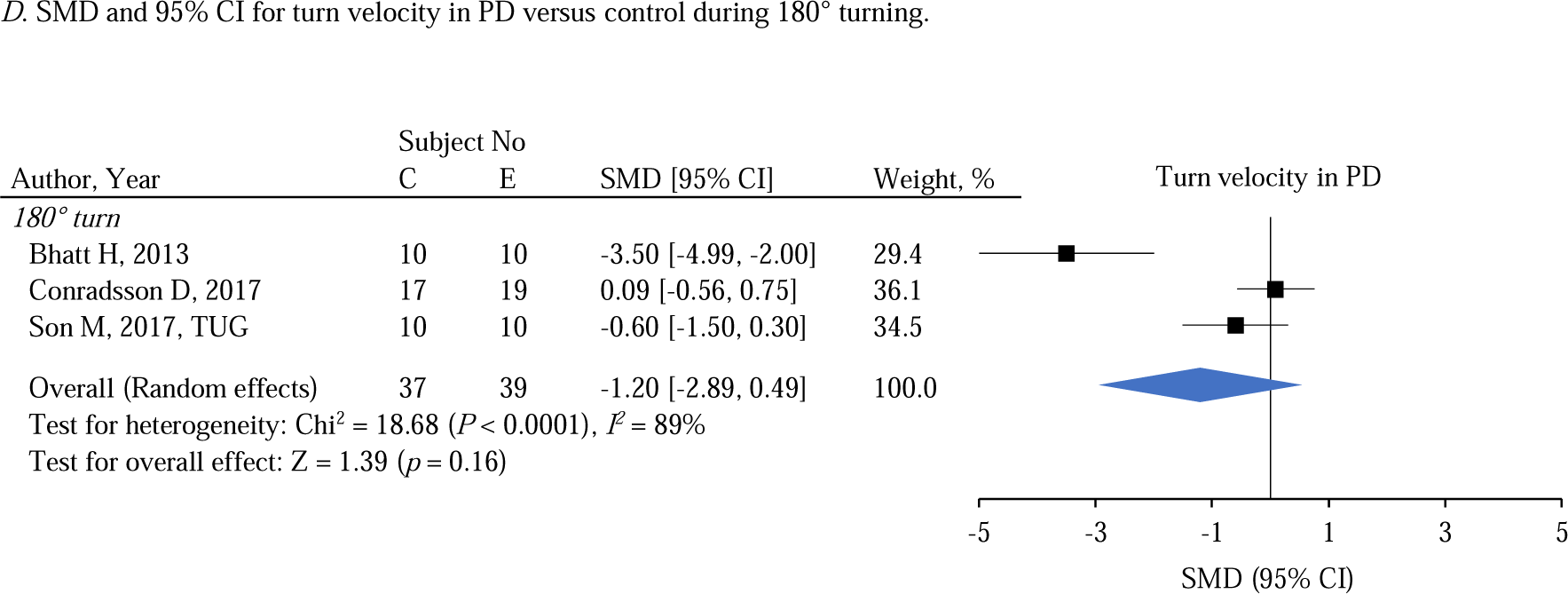
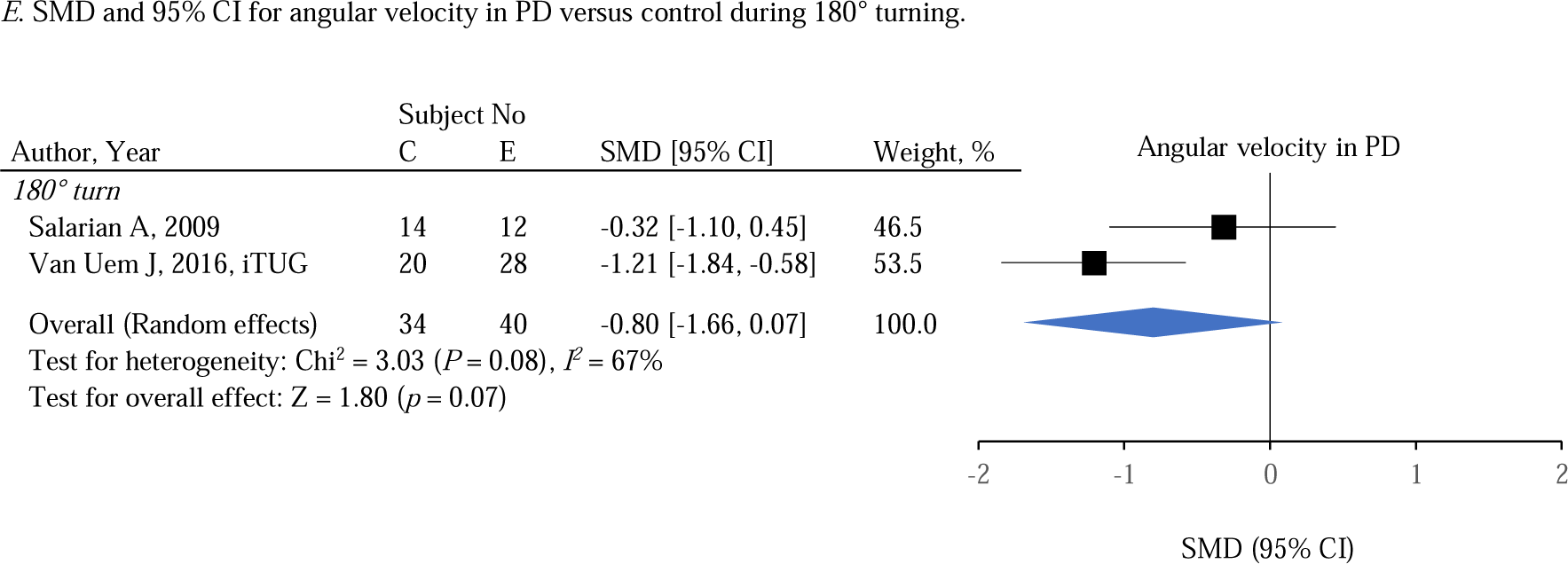
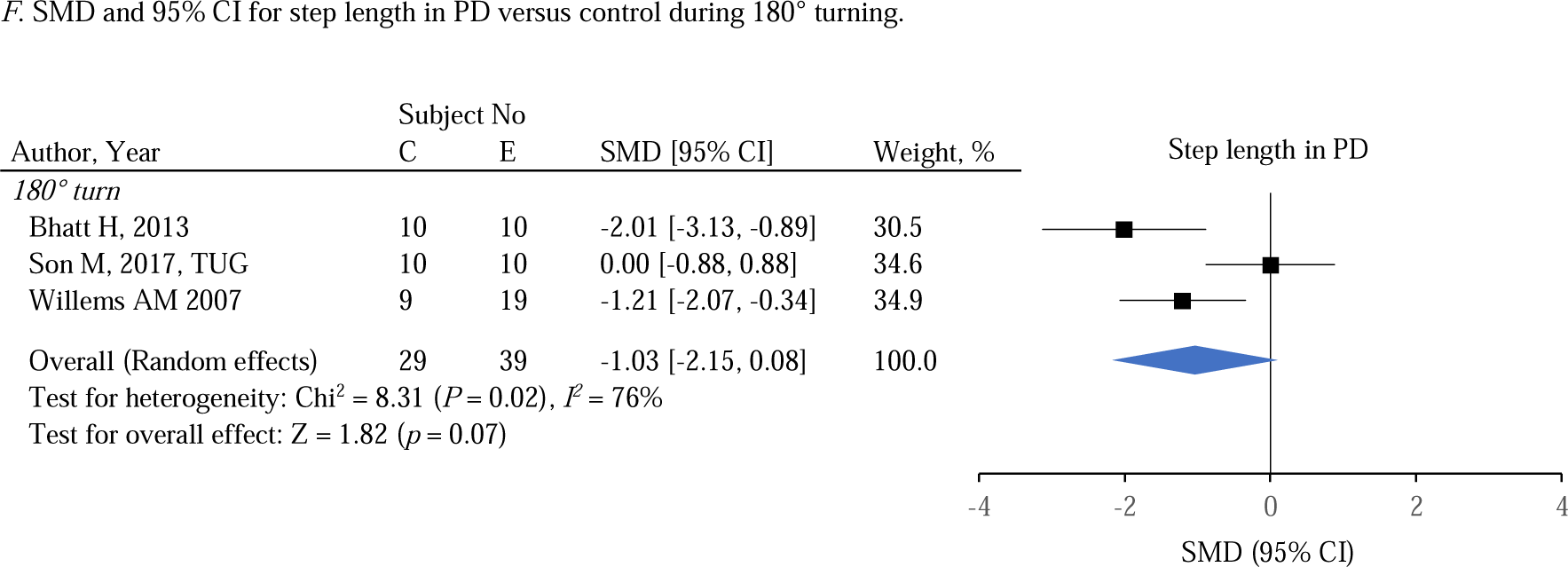
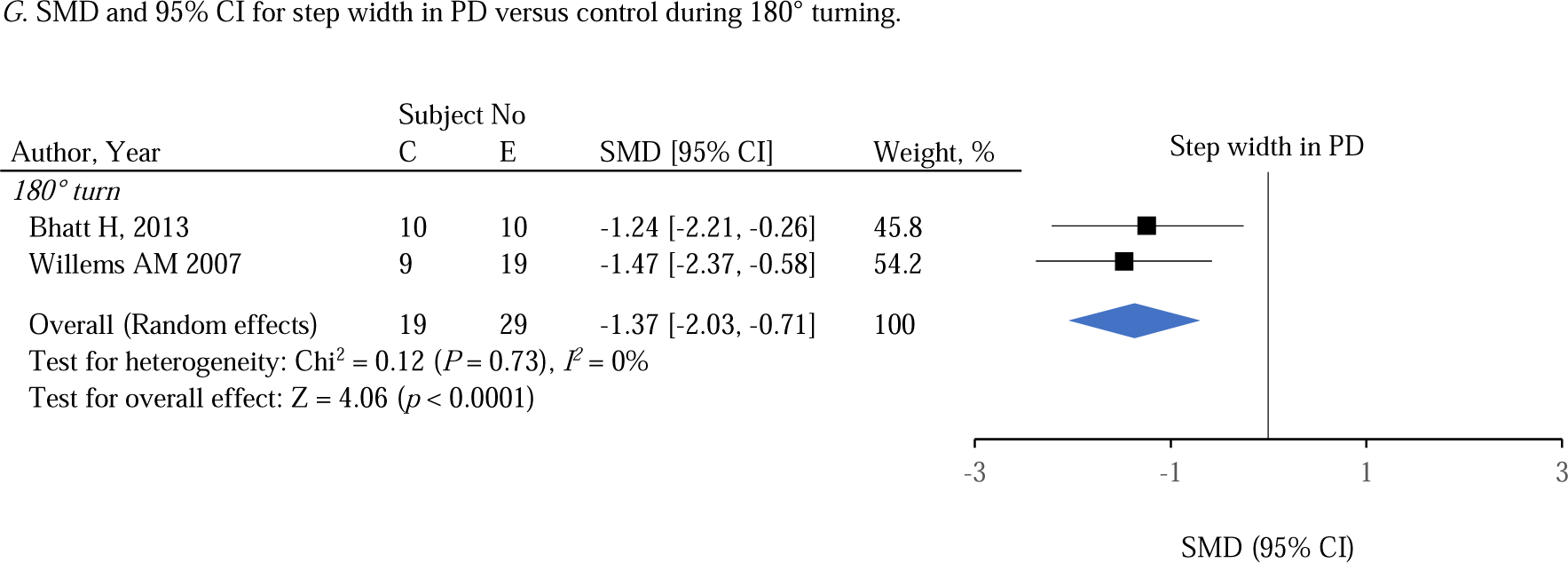
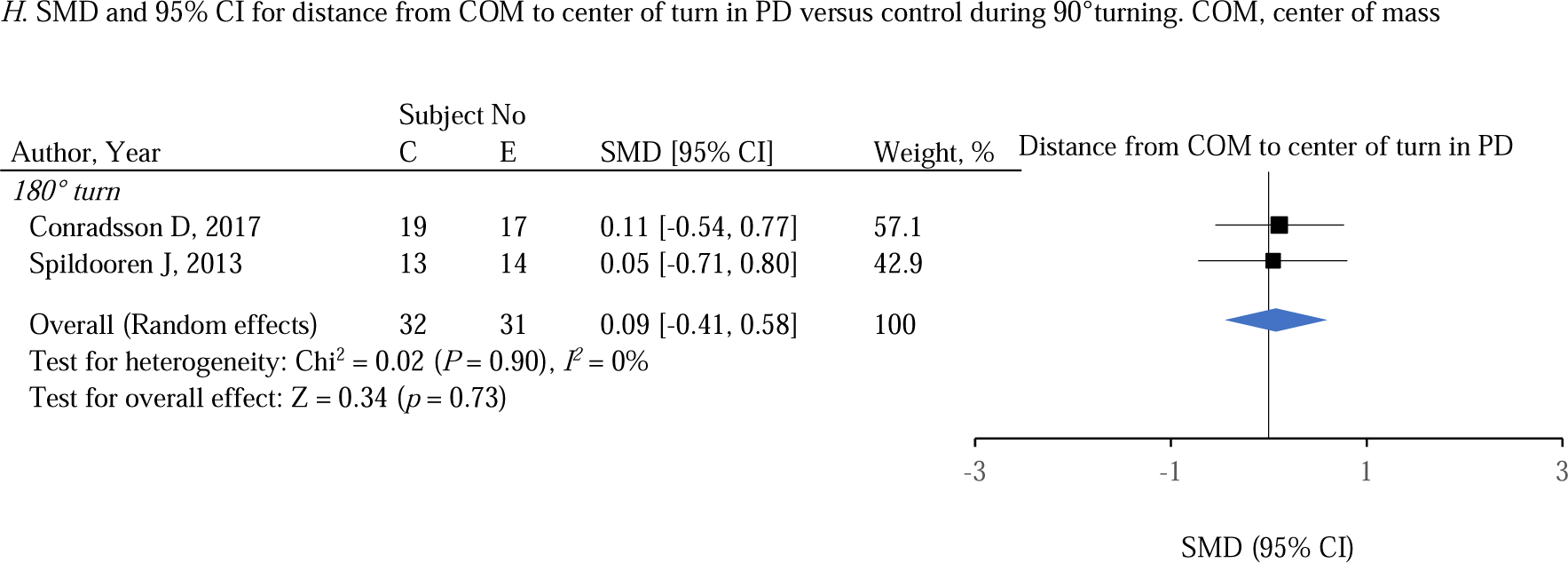
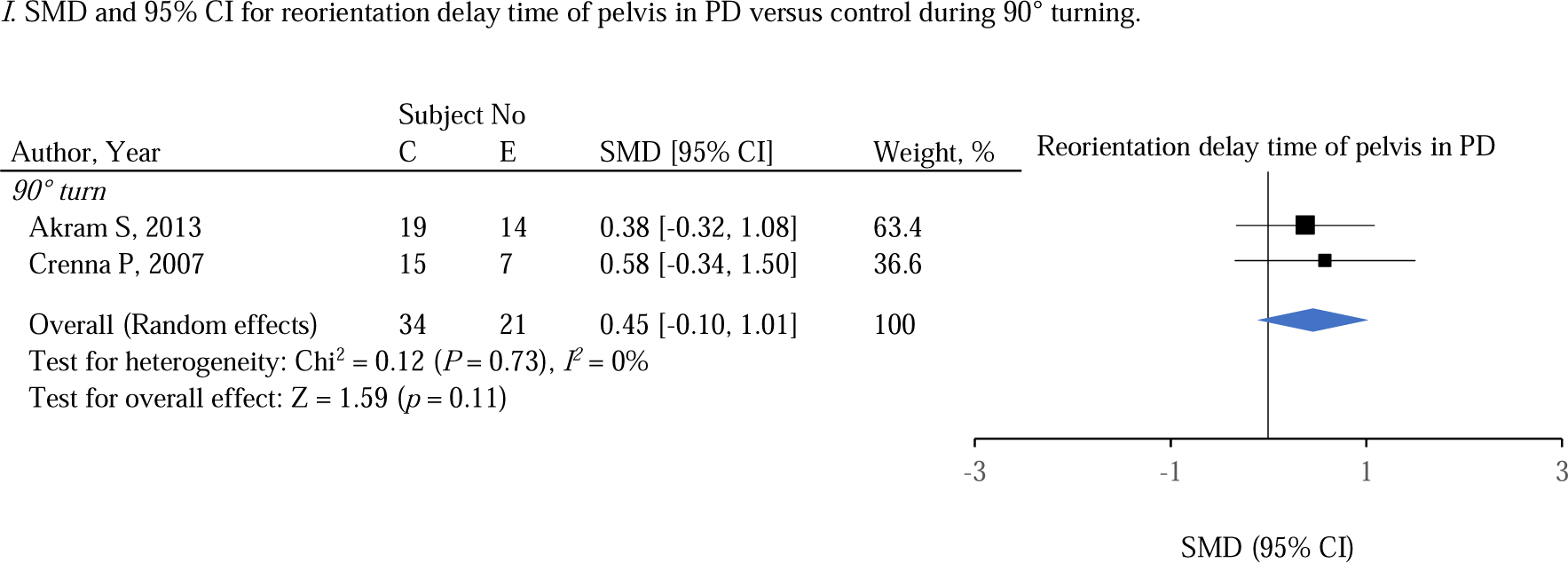
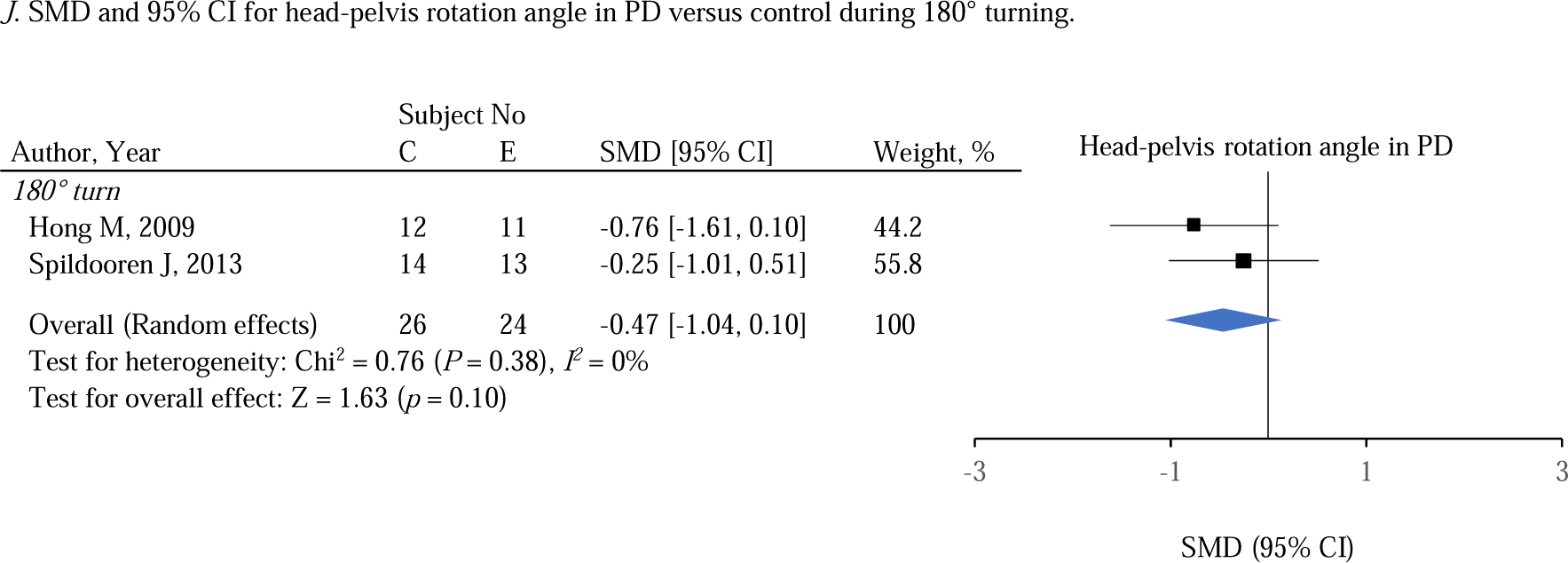

Moreover, kinematic parameters gave us interesting implications although sufficient studies were not gathered for the meta-analysis. The head rotation in patients with PD was also smaller during 90° turning[32, 35]. Son et al. reported a smaller range of motion (ROM) of the hip, knee, ankle, and shoulder, and foot clearance in patients with stroke than controls[62].

The strategy parameter did not show significant trends; however, they were interesting. The distance from COM to the center of turn was slightly larger in PD (**Figure 3*H***[9, 34]), reorientation delay time of pelvis in PD was slower than that of controls (**Figure 3*I***[19, 35]), and head-pelvis rotation angle was smaller than controls (**Figure 3*J***[9, 41]). However, head rotation relative to trunk in PD was smaller than control during 90° turning[32, 35] and 180° turning on the spot[41]. Based on these results with each segment reorientation onset time, controls perform sequential segmental turning from head to pelvis, while patients with PD performs “en bloc” turning which means near-simultaneous rotation of body segments[32, 35, 41, 42, 62].

#### Other diseases

Since the outcomes were not unified in other diseases, there were no parameters available to apply meta-analysis. The individual results are gathered for each disease below.

Turn duration in patients with stroke during 90° turning[40] and 180° turning[11, 30] were longer than those in the control group. Step length and step width during 90° turning[40] and 180° turning[11] were shorter in patients with stroke. Moreover, patients with stroke showed longer stance duration[31], shorter single stance phase and swing phase[11], and slower velocity than controls. The paretic limbs of the stroke group showed lower peak hip extension, knee flexion, ankle dorsiflexion[11], and larger foot clearance while shorter of nonparetic side than controls[30]. Delay time of the head, trunk, and pelvis reorientation and these order of patients with stroke were similar to controls during 180° turning[47], while inverse order of segment reorientation from the pelvis to the head was reported in case of fast walkers’ paretic turn and slower walkers’ non-paretic turn during a 90° turn[45]. The turn type selection ratio was similar to that in the control group[54].

There were no significant differences between the patients with CAI and control groups on ankle inversion angles during 45° step and spin turn movements[44], neither hip nor knee joint angles during 45° step and spin turns[44]. Contrarily, people with CAI performed smaller maximum ankle inversion angle than controls during the 180° turn task[39], as they impaired controlling their ankle joints in situations where they were likely to be injured than those in the control group.

Patients with TKA performed turning at a lower cadence compared to healthy controls[46]. A decreased ROM at the involved side for the stance phases which was caused by the increased peak knee extension at midstance during walking followed by a sidestep was reported[46]. TKA-involved-knee showed a shift towards increased external and decreased internal rotation in axial plane[46]. The magnitude of acceleration on the knee was significantly larger in patients with TKA compared with controls during 90° turning[43].

Regarding CA, patients with ataxia showed a reduced cycle duration[51], lower body rotation values[61], and a large number of steps[61] compared to the control group. In addition, patients with ataxia showed significantly lower values of peak hip flexion, hip extension ROM, peak knee flexion, knee extension ROM, and ankle plantar-flexion ROM than the controls[61]. Patients with CA never performed spin turns for stability while those in the control group often executed spin turns[51].

Premutation carriers with FXTAS showed significantly longer turn duration compared to controls[55]. Patients with HD showed significantly longer turn duration and greater center of gravity sway velocity[56]. Patients with TMA who had partial foot amputation displayed significantly longer TUG times compared to controls[63].

## Discussions

### Disease specific turn characteristics

This study revealed the gap of knowledge regarding the characteristics of turning movement for each disease by comprehensively gathering studies that have been individually conducted for each disease, such as bias in amount of knowledge and disparate methodologies. Nervous system diseases, especially PD was the most investigated in previous studies among nervous system and musculoskeletal diseases dealing with turning tasks.

Among the reviewed studies, the most common parameter was spatiotemporal parameters such as turn duration and the number of steps. These parameters are comparably easy to acquire with simple apparatus such as a stopwatch and video camera. Moreover, strategy parameters especially segmental and turn type subcategories were often reported. The overview of the disease-specific turn characteristics is shown in **Figure 4**. Common features among several diseases were slower movement in PD[9, 25, 29, 32, 35, 41, 48, 49, 52, 59, 60, 62, 64, 66, 68-70], stroke[11, 30, 40], FXTAS[55], HD[56], and TKA[63] than those in the control group, which were reflected in longer turn duration and/or slower velocity. However, only patients with CA[51] and CAI[44, 71] did not show slow movements. Contrarily, the trends of cadence, number of steps, step length, and step width were different for each disease. Patients with CA[51, 61], stroke[11, 40], and PD[29, 70] performed turns with smaller step lengths than those in controls, and this result in a greater number of steps. Different from patients with PD and stroke, patients with CA performed turns with fast steps to walk as rapidly as the individuals in the control group[51]. However, a wider step width is reported as a feature of CA for compensatory instability[51]. Patients with FXTAS performed turn with as a large step length as those in the control group, but the variability was large because of the instability[55]. Few features during turn were confirmed in musculoskeletal diseases discussed in the current study. Especially in patients with CAI, the authors reported a difficulty to distinguish the patient and controls by observing turn task[44, 71]. Characterizing a turn in those musculoskeletal diseases may be difficult. PD is characterized by a large cadence[29]. Although the cadence and turn duration of PD and CA cannot directly be compared, patients with CA are considered to have a higher ability to move forward than PD, since patients with CA perform turns at a similar velocity as controls. Patients with stroke have been reported to have a fewer cadence than controls[11] different from PD. Regarding strategy parameters, the turn type was likely to be disease dependent[51, 54, 67]. The patients with PD[8, 29] and CA[51] tended to avoid spin turn and select multi-step turn while turn type selection in patients with stroke was similar to controls[54]. Spin turn is known as efficient but unstable because the COM move out of the base of support. The high incidence of spin turns in the elderly may be related to the risk of falling[72]. PD and CA patients chose more stable turn type different from patients with stroke. Regarding the segmental parameters, there was a difference between CA, stroke, and PD in terms of segment reorientation timing; patients with CA[51, 61] and stroke[47, 54] were capable of “steering turn” that head, trunk, and pelvis segments were reorienting in order, while PD performed “en bloc”[32, 35, 41, 42, 62] that simultaneous and delayed segments reorientation. These disease-specific movement characteristics may allow for early screening.

**Figure 4.**
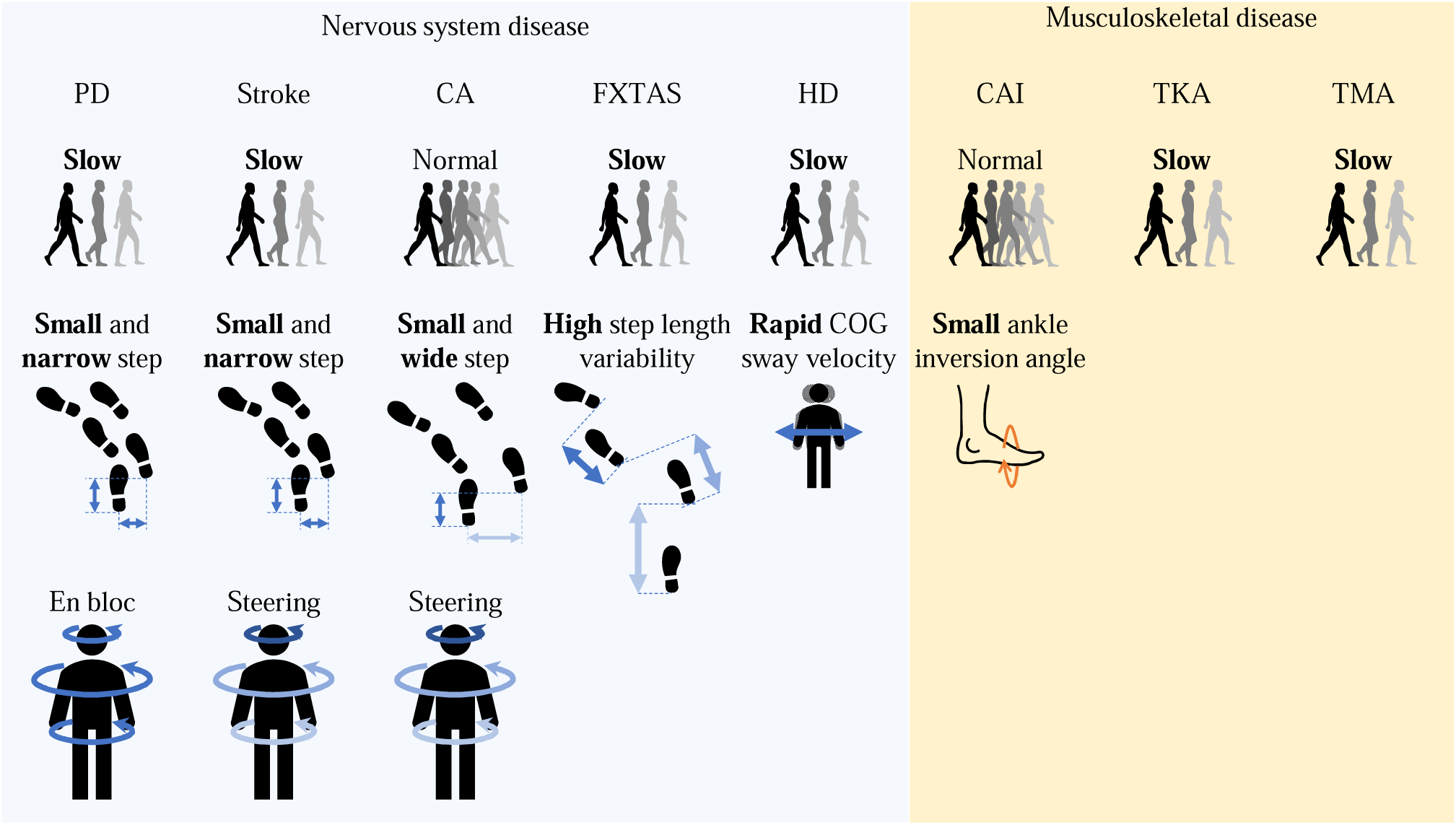
The overview of the disease-specific turn characteristics.

### Characteristics of patients with PD and effect of task variation

PD-specific turn characteristics are explained by slow multi-step and “en bloc” performance. The results of the meta-analysis show a larger number of steps, longer turn duration, slower turn velocity, smaller angular velocity, smaller step length, smaller step width, which are mechanisms of slow and multi-step turn. “En bloc” performance was identified by a smaller head-pelvis rotation angle. The segmental characteristics seem more certain than spatial characteristics as the distance from COM to the center of turn showed no differences with controls.

According to the results of the meta-analysis, the turn duration and number of steps during both 180° turn on the spot and 180° turn showed the same trend, while the differences between patients with PD and controls were larger during 180° turn on the spot than 180° turn. This suggests that the task of walking directly backward on the spot without an approach is more difficult than the task with an approach, such as TUG, and that is easier to detect differences from controls. Turning and turning on the spot are both frequent movements, especially in daily life. Turning on the spot often occurs by a combination of other movements, such as after standing up from a chair or taking something out of a refrigerator or shelf[20]. In addition, there is a high possibility of tight turning like turning on the spot in dead-end spaces such as kitchens, toilets, and closets, where there is not enough space for radius turning. Although no disease-specific differences between turning and turning on the spot have been reported[19, 20], turning on the spot is likely to be more difficult for patients with PD, who presents FOG during gait initiation as well as turning[67, 73].

The number of steps[32, 35, 41, 48, 52, 59, 60, 64, 66, 70], turn duration[9, 48, 49, 59, 60, 64, 68, 70], and step length[29, 62, 70] showed similar trends at 90° and 180°; thus, the same trends can be shown regardless of the angle. However, a similar trend was shown in both angles (90° vs. 180°) in step time[9, 32, 35, 59, 60, 62, 64, 70] which has a little difference between PD and controls.

During the 45° spin turn, longer step length, faster step speed, and longer step time was required than 45° step turn and 90° step turn[67]. This means the high difficulty of spin turn task for PD and turn type has more impact on the movement than turn angle between 45° and 90°.

### Implication on standardization of task protocol and evaluation indicator for future research

The selecting ratio of turn types differs depending on the turning angle. A study of CA showed that the 90° turning was more likely to differ from the control group comparing to the 30° turning because the patients prioritized safety and performed alternative strategies (avoiding spin turns and adopting multi-step turns) during only 90° turning[51]. Regarding PD, patients showed more multi-step turns instead of step turns during 180° turning than 90° and 120° turning[29]. These studies indicate the effect of turn angle on strategy differ between 30° and 90°, and between 90° or 120° and 180°. Further investigation of the effects of continuous angle change is required. Also, for asymmetrical symptoms, such as hemiplegia, the turning direction should be classified into the ipsilateral and contralateral sides. The strategy selection ratio for each task (ipsilateral/contralateral) proceeded in several studies[29, 34, 66] should be more conducted in the future.

Since the spatiotemporal, kinematic, and kinetic parameters change depending on the turn types[18], pivot foot and turning direction should be at least distinguished for analysis. Previously defined turn type classifications were step/spin[8, 18, 34, 54], step/spin/multi-step[29, 51], sideway/twisting/backward/festination/forward/wheeling[8], toward/pivotal/lateral/incremental/delayed onset[66]. Concerning early screening, the classification of few-step turn including step turn and spin turn, and multi-step turn is standard, and it may be desirable to subdivide the classification by disease for more detailed consideration. Although some studies have originally defined step type classification, major classification is a few-step turn including step turn and spin turn and multi-step turn. Only few-step turn classification is often adopted in turn tasks with smaller turn angles including 90° turn, while a multi-step turn is additionally used in most 180° turn. Adamson et al. classified multi-step turn into four types for 180° turn task[8]. This indicates the variety of multi-step turn in PD. Whereas Stack et al. suggested a unique classification of three types of few-step turn in PD for 180° turn on the spot task[66]. This implies that even with the same 180° turn, the step type classification and selection ratio is different between turn on the spot and walking turn.

The major instruction method of the turning location was showing the turn center with some targets such as poles in the study of PD, stroke, FXTAS, and TMA. In this case, the line of sight and the walkable area differ depending on the height of the landmarks, which may affect the movement parameters. Particularly, the reflective markers or tapes were small; hence, the participants could be stepped over. Turning is a complex task that requires not only physical but also cognitive abilities, as we must decide where to start rotating and which leg with during walking. Thereby, the role of the landmarks as indicators of turning position is important for the assessment of patients’ abilities. However, no study has reported the recognition of landmarks, although the gaze direction is assessed with the reorientation of the other segments in several studies[10, 25, 45]. In cases of center location settings, COM distance from the center of rotation and COM trajectory can be evaluated. The COM distance of patients with dementia was shorter than that of healthy controls, while that of patients with distal radius fracture was longer[74]. Patients with PD did not show a certain trend in total COM distance. Conradsson et al. reported shorter in PD compared to controls[34] whereas Bengevoord et al. reported opposite result[27]. Mellone et al. showed that patients with PD performed walking on the route including several corners with shorter distances than controls[53], while Willems et al. reported patients with PD made a bigger turn around an obstacle than controls[70]. Instruction of the foot position using force plates is major in studies of CAI, CA, and TKA. This implies that kinetics are highly important in these diseases. Although force plates are essential for measuring kinetic parameters, it is a concern that they impede observational differences in disease-specific strategies such as positioning against a landmark at the corner. Clarification of the kinetic characteristics of each disease is important; however, it may be useful to focus on other parameters which are easier to acquire from the perspective of early screening. Moreover, the turning task that the timing was specified by audibly signs[51, 61], displays[34, 45, 54], or lightings[39] is more complex because it reflects the effects of cognitive ability, vision, and hearing function.

Speed specification also needs to be mentioned. Although self-selected speed was used in many studies, Van Uem et al. reported that specifying the walking speed as fast as possible for patients with PD made a more significant difference than self-selected velocity[68].

## Conclusion

In the current study, we conducted a systematic review to reveal the characteristics of turning in nervous system and musculoskeletal diseases associated with gait disorder for the early screening of diseases. Our results reveal that the spatiotemporal parameters (number of steps, turn duration, step length, and step width) and strategy parameters (turn type and segment subcategories) tended to show characteristics of gait specific to patients with PD. Subsequent studies are required since turning is a more difficult movement than normal walking[75], and accounts for a high percentage of walking movements[5]. There was a lack of uniformity in movement measurement conditions and parameters in previous studies because the necessary conditions and framework were not established. A uniform protocol having standardized task conditions should be established to understand the trends for each disease for early screening. This review contributes to show the disease-specific characteristic trend beyond multiple diseases for future studies.

## Data Availability

All data produced in the present study are available upon reasonable request to the authors.

## Acknowledgments

This study was supported in part by JSPS KAKENHI Grant Number JP20K13807. This funding source had no role in the study design, in the collection, analysis and interpretation of data; in the writing of the manuscript; and in the decision to submit the manuscript for publication. We thank editage (https://www.editage.jp/) for the English language editing.

## Author contributions

AO: Conceptualization, Data curation, Formal analysis, Funding acquisition, Investigation, Methodology, Software, Validation, Visualization, Roles/Writing - original draft, Writing - review & editing

TT: Data curation, Formal analysis, Investigation, Software, Roles/Writing - original draft, Writing - review & editing

KY: Data curation, Formal analysis, Investigation, Writing - review & editing

HI: Conceptualization, Methodology, Project administration, Resources, Supervision, Writing - review & editing

## Data statement

The datasets used and/or analyzed during the current study are available from the corresponding author on reasonable request.

